# Accuracy of left ventricular mechanical dyssynchrony indices for mechanical characteristics of left bundle branch block using cardiovascular magnetic resonance feature tracking

**DOI:** 10.1101/2022.11.11.22282225

**Authors:** Daniel E. Loewenstein, Björn Wieslander, Einar Heiberg, Jimmy Axelsson, Igor Klem, Robin Nijveldt, Erik B. Schelbert, Peder Sörensson, Andreas Sigfridsson, David G. Strauss, Raymond J. Kim, Brett D. Atwater, Martin Ugander

## Abstract

**Aims:** More than 90% of patients with left bundle branch block (LBBB) and reduced left ventricular (LV) ejection fraction have LV dyssynchrony and a high probability of response to cardiac resynchronization therapy (CRT). A subgroup of patients with non-specific intraventricular conduction delay (IVCD) have a LBBB-like LV activation pattern when studied using invasive mapping and advanced echocardiographic techniques. These patients also frequently benefit from CRT but these patients have proven difficult to identify using ECG criteria. Cardiovascular magnetic resonance (CMR) imaging indices of dyssynchrony may identify patients with IVCD who may benefit from CRT but their relative accuracies for identification of LV dyssynchrony remains unknown. We compared the LV dyssynchrony classification accuracy of two commonly available CMR indices in a study population of patients with severely reduced LV ejection fraction and no scar, and either LBBB or normal conduction (normal QRS duration and axis, controls).

**Methods and results:** In LBBB (n=44) and controls (n=36), using CMR feature tracking circumferential strain, dyssynchrony was quantified as the circumferential uniformity ratio estimate (CURE) and the systolic stretch index (SSI). Deidentified CMR image-data were made publicly available. Both CURE and SSI quantified more severe dyssynchrony in LBBB compared to controls (p<0.001 for both). SSI more frequently discriminated LBBB and normal conduction LV activation patterns than CURE (area under the receiver operating characteristics curve [95% confidence interval] 0.98 [0.95-1.00] for SSI vs 0.77 [0.66-0.86] for CURE, p<0.001).

**Conclusions:** SSI is superior to CURE for discriminating synchronous and dyssynchronous LV activation and should be further studied in the setting of non-LBBB conduction abnormalities.

## 1. Introduction

Cardiac resynchronization therapy (CRT) has shown major favorable effects for the treatment of patients with heart failure (HF), severe LV dysfunction, and prolonged QRS duration. The COMPANION, REVERSE, and MADIT-CRT trials demonstrated that patients with LBBB derived greater benefit from CRT compared to those with other conduction patterns [1– 3]. Furthermore, strict LBBB, as defined by Strauss, et al., has to an even greater extent been found associated with positive CRT response and also identifies patients with a “super-response” to CRT [4], when compared to LBBB defined by conventional criteria [5, 6]. This is likely a result of greater specificity for the strict LBBB criteria for identifying clinically significant LV dyssynchrony.

Non-specific intraventricular conduction delay (IVCD) is the second most prevalent conduction abnormality amongst patients receiving CRT [7, 8]. IVCD is often grouped together with right bundle branch block (RBBB), atypical LBBB, and atypical RBBB as “non-LBBB”. In non-LBBB, no overall benefit of CRT has previously been shown [9, 10]. Consequently, this grouping has recently been contested in a large patient-level meta-analysis including several pivotal CRT trials [8]. In that study, patients with wide QRS (>150ms) and either LBBB or IVCD had similar effect of CRT in reducing heart failure hospitalization or death [8].

Considering the heterogeneity in the “non-LBBB” group of patients, predicting CRT response based on QRS duration and ECG morphology in isolation could be too simplistic an approach [11]. CMR measures of dyssynchrony may be useful in identifying those patients without LBBB or normal conduction who have mechanical dyssynchrony that may benefit from CRT. CMR studies of regional mechanics might help identify subgroups of patients with LBBB-like characteristics. An index sensitive to the specific mechanical characteristics of strict LBBB and its associated high CRT response rate would potentially facilitate an assessment of dyssynchrony in patients without strict LBBB. Therefore, the aim of the current study was to identify such an index by comparing the discriminatory ability of two currently used methods of CMR dyssynchrony measurement to distinguish patients with strict LBBB from patients with normal conduction. We also sought to control for other myocardial abnormalities that can impact the electro-mechanical association by only including subjects with left ventricular ejection fraction (LVEF) ≤ 35%, and no LV myocardial scar assessed by CMR late gadolinium enhancement (LGE).

## 2. Methods

This is an observational case-control study where patients were retrospectively identified by cross-referencing the CMR and electrocardiography (ECG) databases from three centers (Duke University Medical Center, NC, USA; Pittsburgh University Medical Center, PA, USA; and Karolinska University Hospital, Stockholm, Sweden). The study was approved by the local human subject research ethics committee at each site, and all subjects either provided written informed consent or were included following a retrospective waiver of informed consent provided by the local ethics committee.

### 2.1. Subject selection

Subjects considered for inclusion in the present study had a LV ejection fraction *≤*35%, no scar by CMR late gadolinium enhancement (LGE), CMR cine images in a LV short-axis stack, and either normal ECG QRS duration (*<*120 ms) and frontal plane electrical axis (-30 to +90 degrees, controls, n=36), or LBBB (n=44) defined by Strauss’ strict ECG criteria, defined as a terminal negative deflection in lead *V*_1_ and *V*_2_ (QS or rS configuration), a QRS duration ≥ 140 ms for men and ≥ 130 ms for women, and the presence of mid-QRS notching or slurring in ≥ 2 of leads *V*_1_, *V*_2_, *V*_5_, *V*_6_, I and aVL [4]. Subjects were excluded if they had a history of congenital heart disease, CMR evidence of myocardial storage disease, atrial fibrillation, prior openheart surgery, or LV septal wall flattening indicative of clinically significant pulmonary hypertension. The following baseline characteristics were collected: age, sex, height, weight, body surface area (BSA), body mass index (BMI), and CMR measures of LV volumes, function, and mass. Among the patients who met the inclusion criteria (n=87), patients were excluded due to having Takotsubo cardiomyopathy (n=1), atrial fibrillation discovered at time of feature tracking analysis (n=1), or missing or insufficient number of diagnostic quality CMR cine images (n=5). As a result, the final study group included 80 patients.

### 2.2. CMR image acquisition

All imaging was performed with clinically available scanners at the respective centers. Scanners included 3T (Siemens Verio, Erlangen, Germany) and 1.5T systems (Siemens Avanto, Espree or Aera, Erlangen, Germany, or Philips Intera, Best, the Netherlands), all using ECG gating and phased-array receiver coils. Typical acquisition parameters for cine images were: repetition time 44 ms, echo time 1.2 ms, flip angle 60 degrees, matrix 190 x 190, slice thickness 6 mm, and temporal resolution 24 frames per cardiac cycle. Clinical reports of cardiac viability assessment were reviewed for mention of any myocardial scar by LGE.

### 2.3. Image and strain analysis

Cine CMR images exported for offline myocardial strain analysis performed by an observer using commercially available software for CMR feature tracking (Segment version 3.2 R8757) Medviso, Lund, Sweden) [12, 13]. All analysis was performed blinded to ECG classification. Endocardial and epicardial borders, excluding papillary muscles and trabeculations, were manually delineated in the end-diastolic reference timeframe. The end-diastolic reference timeframe was set to the timeframe immediately following halting of the circumferential expansion and longitudinal lengthening of the LV during late diastole as viewed in a three chamber, long-axis, and short-axis slice. This was due to observation that there was often a delay in the closure of the mitral valve, which otherwise has commonly been used to define end-diastole. The delineation was performed in a single midventricular short-axis slice. A non-rigid elastic registration strategy was used by the software to measure myocardial strain over time. For regional strain assessment, the area encompassed by the endo- and epicardial borders was segmented into regions of interest according six segments of the American Heart Association 17-segment model. In short-axis images, the location of regional segments was determined using an angle relative to the right ventricular anterior insertion point. Circumferential strain was evaluated from the Lagrangian strain tensor between adjacent points. Mechanical dyssynchrony was quantified using the circumferential uniformity ratio estimate (CURE) [14–16] and the systolic stretch index (SSI) [17, 18]. In short, CURE is derived from Fourier transformation of the spatial distribution of strain from myocardial segments averaged over the number of short axis slices. CURE is then calculated as

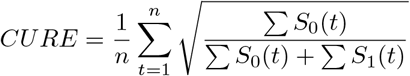

where *S*_0_ is the zero order, and *S*_1_ is the first order term in the fourier transformation, and *n* is the number of timeframes covering the cardiac cycle. CURE ranges between 0 (perfect dyssynchrony) and 1 (perfect synchrony). SSI was originally developed through computer simulations [18], and later presented in a slightly simplified version for use in echocardiography [17] and is calculated as the sum of LV lateral wall systolic pre-stretch (SPS) and septal rebound stretch (SRS).

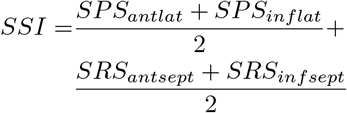

Systolic pre-stretch is defined as the sum of LV lateral wall stretch before aortic valve opening, averaged over the anterolateral and inferolateral segment. Septal rebound stretch is defined as the sum of septal stretch following early systolic shortening and before aortic valve closure, averaged over the anteroseptal and inferoseptal segment.

### 2.4. Statistical analysis

Categorical data are reported as number and percentages. Continuous variables are reported as median [interquartile range]. For continuous variables, groups were compared using the Wilcoxon signed-rank test. Bivariate correlation was examined by Spearman’s *ρ* correlation coefficient. Univariable logistic regression models with LBBB status as the dependent variable were fitted separately for the two dyssynchrony indices. Specificity, sensitivity, discriminatory performance, and cut-off values were derived from receiver-operating characteristics (ROC) analysis using the Youden’s index. Multivariable linear regression models fit separately for each dyssynchrony index in LBBB, and controls respectively, were used to test for associations with covariates age, LV end-diastolic volume index (LVEDVI), LV mass index (LVMI), and sex, indicating any need for covariate-adjusted, or covariate-specific ROC curves. Nonlinearities were entertained by use of restricted cubic splines, as were interactions between LVEDVI, and LVMI with sex, respectively. Areas under the paired ROC curves were compared using nonparametric stratified bootstrapping. Bootstrapped CIs were derived from 4000 replicates and calculated using the percentile method. A two-sided *p <* 0.05 was considered statistically significant. Data processing and statistical analysis were performed in the R statistical programming environment 4.1.0 [19], using package dplyr 1.0.7 [20] for data transformation, ggplot2 3.3.5 [21] for graphical visualizations, pROC 1.17.0.1 [22] for ROC analysis, rms 6.2.0 for regression modeling, and knitr 1.33 [23] for reproducible documentation.

## 3. Results

### 3.1. Subject characteristics

The characteristics of patients included in the study (n=80, 56% female) are presented in Table 1. Characteristics were similar in the two groups except for older age and greater LV mass in subjects with LBBB.

**Table 1.** Data are presented as median [interquartile range] or numerator (percentage). LVEDV = left ventricular end-diastolic volume, LVEDVI = left ventricular end-diastolic volume index, LVM = left ventricular mass, LVMI = left ventricular mass index, LVEF = left ventricular ejection fraction, BMI = body mass index, BSA = body surface area. Tests used: ^1^Wilcoxon test; ^2^Pearson test.

|  | Control<br>*N* = 36 | LBBB<br>*N* = 44 | P value |
| --- | --- | --- | --- |
| Age, years | 48 [36-60] | 64 [59-69] | $p < 0.001^1$ |
| Male Sex, n (%) | 16 (44) | 19 (43) | $p = 0.91^2$ |
| Height, cm | 169 [160-177] | 169 [157-177] | $p = 0.552^1$ |
| Weight, kg | 84.7 [72.1-95.3] | 79.5 [63.8-89.0] | $p = 0.205^1$ |
| BMI, kg/m<sup>2</sup> | 27.3 [25.4-34.2] | 26.7 [24.1-29.4] | $p = 0.282^1$ |
| BSA, m<sup>2</sup> | 1.9 [1.8-2.1] | 1.9 [1.6-2.1] | $p = 0.308^1$ |
| EDV, mL | 254 [164-307] | 255 [210-291] | $p = 0.719^1$ |
| EDVI, mL/m<sup>2</sup> | 123 [101-150] | 127 [114-154] | $p = 0.301^1$ |
| LVEF, % | 25 [21-30] | 27 [24-32] | $p = 0.43^1$ |
| LVM, g | 183 [156-223] | 159 [131-184] | $p = 0.001^1$ |
| LVMI, g/m<sup>2</sup> | 100 [82-117] | 85 [67-96] | $p < 0.001^1$ |
| QRS duration, ms | 90 [84-101] | 158 [150-170] | $p < 0.001^1$ |

### 3.2. Dyssynchrony measurements

Both CURE and SSI showed group differences between LBBB and controls (Figure 1). Consistent with a greater amount of mechanical dyssynchrony in LBBB, CURE was lower in LBBB compared to controls (0.63 [0.54-0.75] vs 0.79 [0.69-0.86], p<0.001), and SSI was higher in LBBB compared to controls (9.4 [7.4-12.7] vs 2.2 [1.2-3.6], p<0.001). Compared to CURE, SSI had a greater area under the ROC curve for detecting mechanical dyssynchrony associated with strict LBBB (0.98 [95% CI: 0.95-1.00] vs 0.77 [95% CI: 0.66-0.86], p< 0.001, Figure 2), and this corresponded to a higher sensitivity and specificity for SSI compared to CURE (Figure 3). The odds ratio (OR) for identifying LBBB for SSI was 3.40 [95% CI: 1.84-6.28] per 1 percentage unit increase in SSI value, and for CURE was 2.16 [95% CI: 1.44-3.24] per 0.10 decrease in CURE value. In evaluating the need for covariate-adjusted and/or covariate-specific ROC curves, linear regression models were used to test the association between dyssynchrony indices and covariates: age,

**Figure 1.**
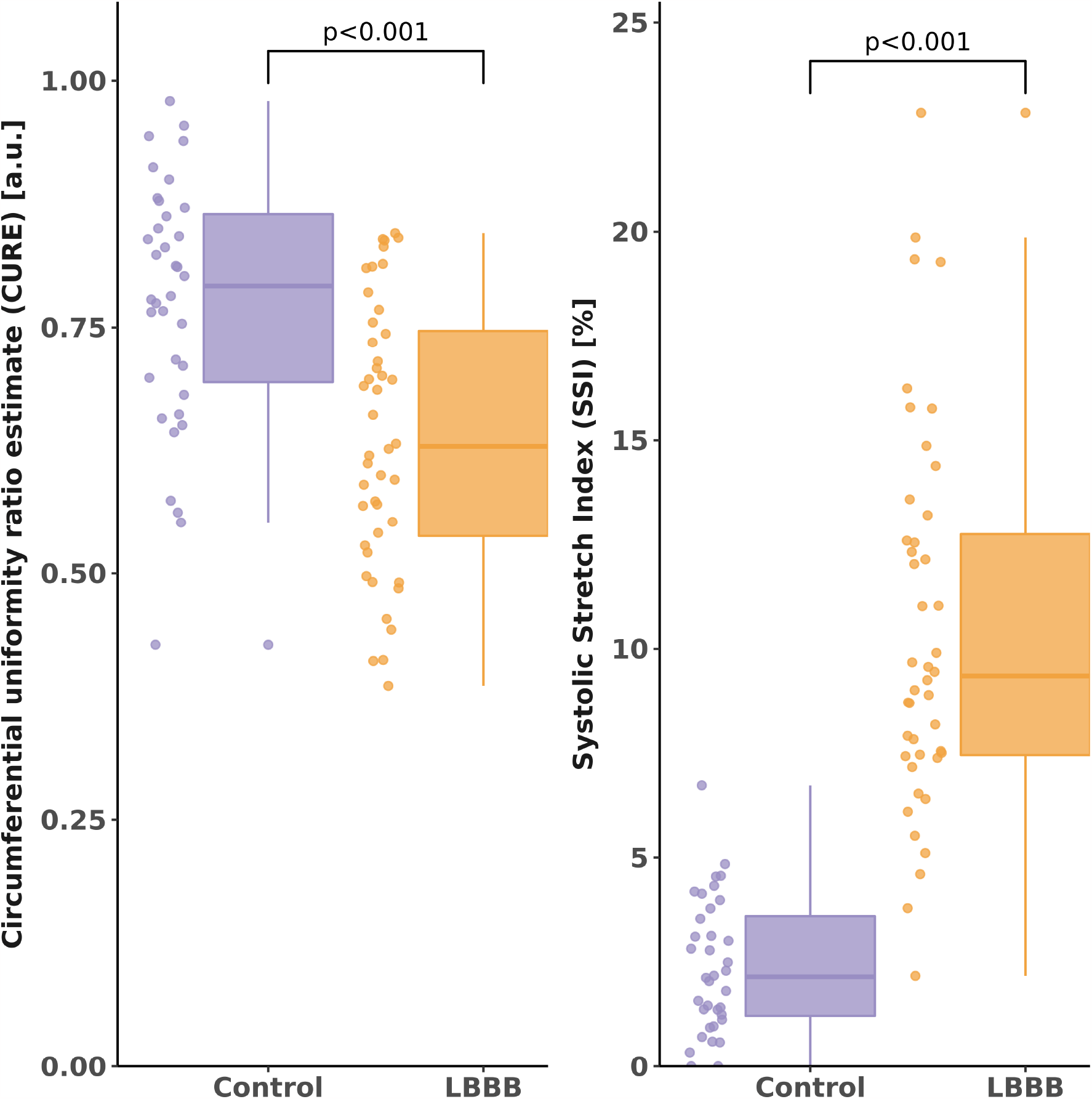
Values for the circumferential uniformity ratio estimate (CURE) and the systolic stretch index (SSI) for left bundle branch block (LBBB) and controls. The box and whisker plots show the median (horizontal line), interquartile range (box), and data points dyssynchrony (lower CURE, higher SSI) in LBBB compared to control. CURE is more homogenously distributed between groups compared to SSI. within 1.5*×* interquartiles ranges of the first and third quartile, respectively (whiskers). Note, there is more pronounced mechanical

**Figure 2.**
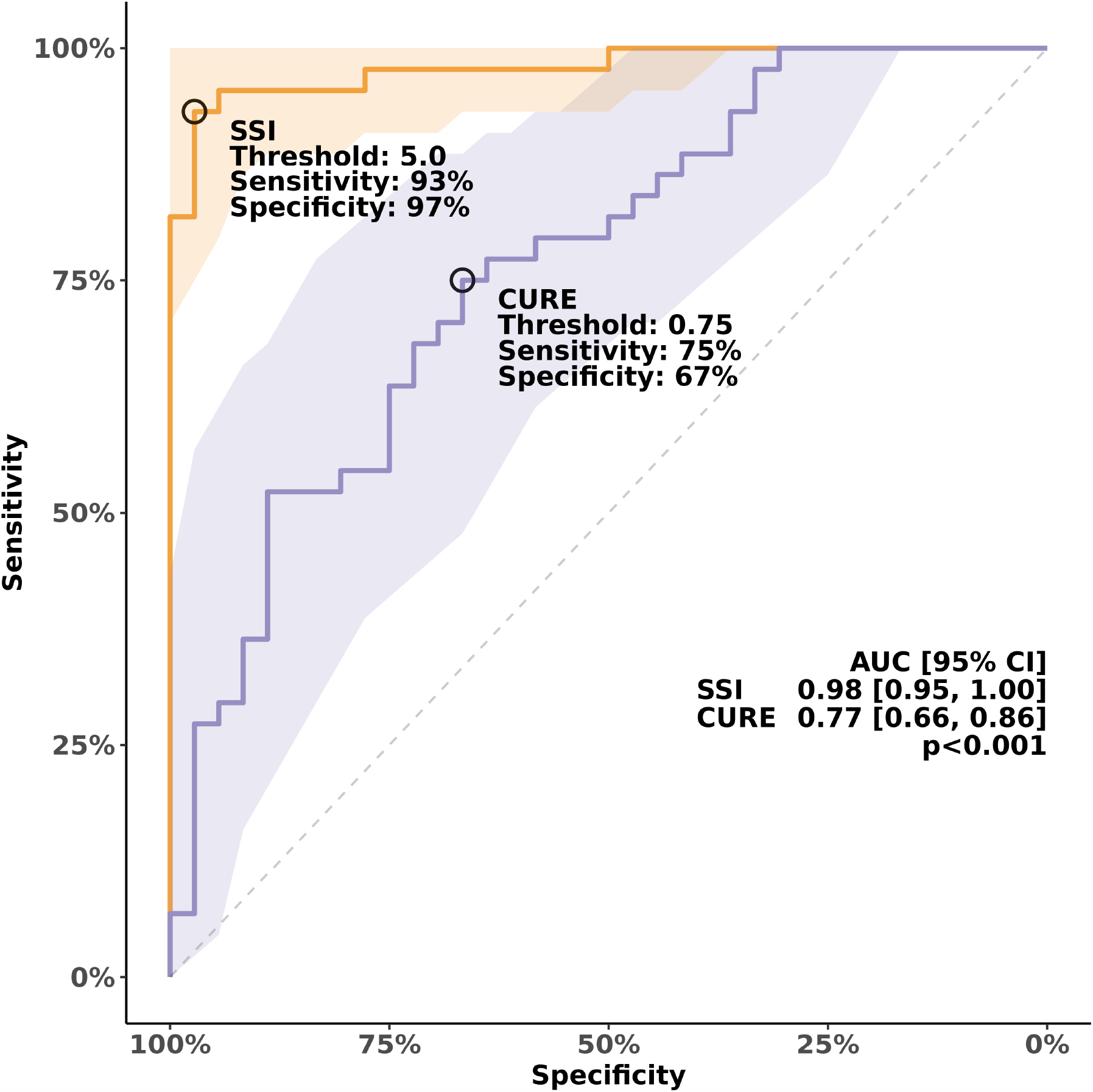
Receiver operating characteristics (ROC) for univariable logistic regression models to differentiate between left bundle branch block (LBBB) and controls using the circumferential uniformity ratio estimate (CURE) and the systolic stretch index (SSI), respectively. Better discriminatory ability for LBBB is seen for SSI compared to CURE. AUC = area under the ROC curve, CI = confidence interval.

**Figure 3.**
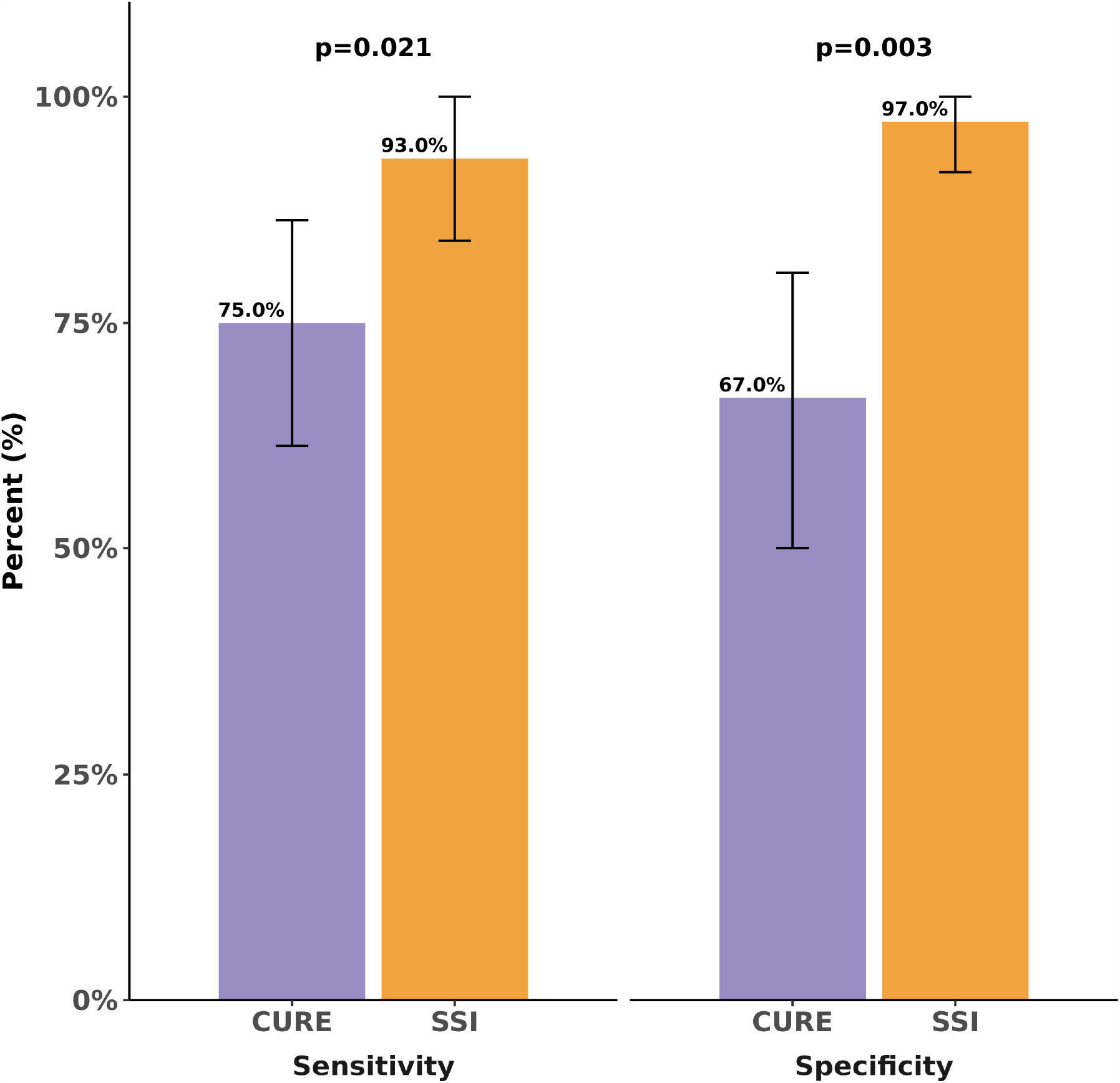
Bar plot showing sensitivity and specificity for detecting left bundle branch block using the systolic stretch index (SSI) and the circumferential uniformity ratio estimate (CURE), respectively. Error bars denote 95% confidence intervals.

LVEDVI, LVMI, and sex, allowing for interactions between LVMI and LVEDVI, with sex, respectively. No evidence was found in support of an association between dyssynchrony indices and covariates in controls, or between CURE and covariates in LBBB. Furthermore, neither CURE nor SSI associated with QRS duration in either LBBB or controls (p ≥ 0.4 for all). There was an association between SSI and age, keeping other covariates fixed, in LBBB. This suggests that the discriminatory ability of SSI might vary with respect to age. For purposes of clarity, the unadjusted ROC curve is presented.

### 3.3. CMR image availability

All CMR images analyzed as part of the current study are made available online (doi: 10.6084/m9.figshare.15155596). Among the individual CMR exams (n=80) the majority were performed on a Siemens scanner (n=75) and only a few on a Philips scanner (n=5), all contain a cine short-axis stack, while most also contain cine 2-chamber (n=78), 3-chamber (n=79), and 4-chamber images (n=79). As per the inclusion criteria all patients were verified to be free of myocardial scar by LGE CMR.

## 4. Discussion

The main finding of the study is that the ability to discriminate between LBBB and normal QRS duration among patients with severely reduced LV ejection fraction and no scar was fair for CURE and excellent for SSI. This highlights that, when developing and evaluating indices aimed at accurately identifying mechanical dyssynchrony amenable to CRT, it is important to evaluate the performance of a proposed index both in patients with LBBB, and in comparison to control subjects with normal QRS duration.

The lack of response and even harmful effects of CRT when implanted in patients with narrow QRS complexes indicates that CRT requires an electrical substrate [24, 25]. Findings that LBBB patients derive greater benefit from CRT compared to those without support that LBBB is this electrical substrate [1–3]. Response to CRT can partly be explained by correction of the discoordinated contraction of myocardial wall segments, a consequence of abnormal electrical activation. However, not all patients with LBBB by conventional electrocardiographic criteria have complete LBBB [26, 27], and mechanical dyssynchrony is not uniquely associated with electrical dyssynchrony [28, 29]. For example, focal LV myocardial scarring is also known to cause abnormalities in regional myocardial mechanics [29]. Approximately one third of patients with LBBB by conventional criteria do not have strict LBBB [30]. Consequently, Strauss, et al, proposed more strict criteria for LBBB [4]. Increased rate of CRT response has been found when using this strict definition of LBBB [5, 6, 31], and strict LBBB is associated with greater mechanical dyssynchrony than non-strict LBBB [32]. Mechanical dyssynchrony per se might therefore not be of primary interest in predicting CRT response but instead focus should be on identification and quantification of the mechanical dyssynchrony pattern associated with an abnormal electrical activation as seen in complete LBBB. Importantly, the current study evaluates the discriminatory ability of two recently proposed mechanical dyssynchrony indices for the mechanical dyssynchrony pattern associated with strict LBBB compared to patients with equally reduced ejection fraction but normal QRS, while controlling for confounding factors such as scar. QRS duration cannot accurately characterize the spectrum of conduction abnormalities, and so it seems unlikely that any singular mechanical dyssynchrony index will be able to capture the full spectrum of variation inherent to dyssynchronous ventricular contraction. While both CURE and SSI capture differences in the pattern of discoordination on a group level, we found SSI superior to CURE with regards to the ability to differentiate between LBBB and controls. Our results suggest that the incremental mechanical dyssynchrony component associated with LBBB is better characterized by quantification of the absolute extent of stretch during the opposing directions of movement in the septum and lateral wall.

SSI was developed by Lumens et al. [18] in an attempt to characterize the electromechanical substrate that may respond to CRT. They used a computational model to simulate electromechanical and nonelectrical substrates of mechanical dyssynchrony and identified strain characteristics specific for the different substrates of mechanical dyssynchrony. In a study of patients enrolled in the Adaptive CRT trial, it was found that SSI by echocardiography was independently associated with CRT outcome, adjusting for QRS morphology, QRS duration, sex, heart failure etiology, and treatment with angiotensin-converting enzyme inhibitors/angiotensin II receptor blockers [17]. However, whether SSI has added prognostic value over strict LBBB morphology is still unknown. It remains to be explored whether SSI can be used to identify non-LBBB patients that may be suitable for CRT, and such studies are justified.

CURE was first evaluated in a canine model of heart failure and LBBB conduction delay [14, 15]. It was found that biventricular pacing leads to greater synchrony (increased CURE), improved global function, and that circumferential dyssynchrony indices had greater dynamic range when compared to longitudinal indices [14]. Importantly, CURE was found sensitive to regionally clustered dyssynchrony [14]. Regionally clustered dyssynchrony might show equal variance as dispersed dyssynchrony when compared to variance-based dyssynchrony measures, although with very different effects on cardiac mechanics [14]. While CURE can be considered a more general measure of dyssynchrony, it has been found to be predictive of CRT response in clinical cohorts [16, 33, 34]. An advantage of CURE over commonly used time-to-peak based indices is that CURE utilizes information of the full cardiac cycle. Additionally, considering that CURE is derived from the relative positions of included segments and less so on their absolute value of strain, CURE is theoretically less sensitive to inter-vendor variations of strain measurements. However, despite these theoretical advantages of using CURE, the current study shows that CURE had a modest performance in identifying LBBB specific mechanical dyssynchrony.

The association between mechanical dyssynchrony, quantified as the systolic dyssynchrony index, and myocardial scar has been studied in patients with systolic heart failure [35]. They conclude that 25% of patients with narrow QRS (*<*130 ms) presented with mechanical dyssynchrony, despite no difference in scar burden compared to narrow QRS patients without mechanical dyssynchrony. Those findings suggest that mechanical dyssynchrony in such patients might be secondary to myocardial scar rather than electrical dyssynchrony. There is no general agreement upon the definition of mechanical dyssynchrony, and the difference in vendor software for strain measurements limits straightforward comparisons between studies. CURE has been shown to identify a greater magnitude of dyssynchrony (lower CURE values) in patients with non-ischemic cardiomyopathy compared to healthy controls (0.79 ± 0.14 vs 0.97 ± 0.02) [36]. In a different study, CURE in healthy controls volunteers has been shown to be 0.87 ± 0.07 [37]. The current study shows that patients with severely reduced LVEF and normal QRS duration have some degree of mechanical dyssynchrony even in the absence of scar (median CURE 0.79). This would suggest that other factors beyond scar and LBBB contribute to mechanical dyssynchrony detected by CURE. Such factors may include variations in pre-load and/or afterload, and regional wall motion abnormalities due to chronic ischemia or other non-ischemic cardiomyopathies that impair contractile function without causing myocardial scar.

While CURE and SSI both displayed group differences between LBBB and controls, the current study found no evidence in support of a relationship between either CURE or SSI, and QRS duration within LBBB and control groups, respectively. CURE and QRS duration have previously been found to be modestly correlated (*r* = -0.58; *p<*0.001) in a cohort (*n* = 43) of cardiomyopathy patients with similar reductions in ejection fraction and prolongation of QRS duration, though QRS morphology was not reported [16]. However, when only evaluating the correlation between CURE and QRS duration in those patients referred for CRT (*n* = 20) those authors found that the evidence did not support any correlation (*r* = −0.40; *p* = 0.08) [16]. The apparent lack of correlation between dyssynchrony and QRS duration is of interest considering that current guidelines are still unclear regarding the group of patients with intermediate QRS width (QRS 120-149ms). These exploratory results add to the notion that there is a complex relationship between electrical and mechanical dyssynchrony.

### 4.1. Image availability

The current study shows that patients with severely reduced ejection fraction and normal QRS duration have a baseline level of mechanical dyssynchrony that is not attributable to myocardial scarring or prolonged depolarization of the myocardium. Consequently, when developing an index of mechanical dyssynchrony, specificity for its intended use should be of interest. To our knowledge, no study to date has included patients free of myocardial scar, with a normal QRS duration, and with severely reduced ejection fraction when comparing or developing indices of mechanical dyssynchrony. In order to facilitate future research where baseline mechanical dyssynchrony is accounted for, the images from the current study are made available online. See the section ‘Availability of data and materials’ below.

### 4.2. Limitations

Identification of the time point for aortic valve opening, and aortic valve closure was performed by visual assessment of CMR cine images, and the accuracy of this assessment is limited by the temporal resolution of CMR images. However, any variations in accuracy would equally affect the analysis of both patient groups, and hence should not have a major effect on the overall results. The software used for strain analysis reports segmental strain measurements according to the AHA 17-segment model. Hence, CURE calculation was limited to Fourier transformation applied to six individual myocardial segments in the midventricular short-axis slice. The impact of the spatial resolution of measurement in CURE quantification has not previously been reported. However, it cannot be excluded that quantification of CURE using higher spatial resolution could potentially influence the results.

## 5. Conclusions

SSI was superior to CURE regarding the ability to discriminate between strict LBBB and normal QRS duration among patients with severely reduced ejection fraction and no scar. SSI merits further evaluation for detecting dyssynchrony in IVCD. The amount of dyssynchrony in patients with no scar, and severely reduced ejection fraction, needs to be taken into account when developing and evaluating indices aimed at accurately identifying mechanical dyssynchrony amenable to CRT. Such an evaluation should preferably include control subjects with normal QRS duration, severely reduced ejection fraction, and the absence of myocardial scar, and imaging data from such patients is provided for public use.

## Data Availability

The datasets generated and/or analyzed during the current study, as well as code needed to reproduce all aspects of the current study, are available in the Figshare repository,
https://doi.org/10.6084/m9.figshare.15155596.
The most recent version of the analysis code is available in the Github repository, https://github.com/dloewenstein/dillacs-study.

https://github.com/dloewenstein/dillacs-study

## Appendix

## Acknowledgments

Not applicable.

## Funding

This work was funded in part by grants to MU from New South Wales Health, Heart Research Australia, University of Sydney, Swedish Research Council, Swedish Heart and Lung Foundation, Stockholm County Council, and Karolinska Institutet.

### Abbreviations

AUC: Area under the curve
BMI: Body Mass Index
BSA: Body Surface Area
CI: Confidence Interval
CMR: Cardiovascular Magnetic Resonance
CRT: Cardiac Resynchronization Therapy
CURE: Circumferential Uniformity Ratio Estimate
ECG: Electrocardiography
HF: Heart Failure
LBBB: Left Bundle Branch Block
LGE: Late Gadolinium Enhancement
LV: Left ventricular
LVEDV: Left Ventricular End-Diastolic Volume
LVEDVI: Left Ventricular End-Diastolic Volume Index
LVEF: Left Ventricular Ejection Fraction
LVM: Left Ventricular Mass
LVMI: Left Ventricular Mass Index
OR: Odds Ratio
RBBB: Right Bundle Branch Block
ROC: Receiver Operating Characteristics
SPS: Systolic Pre-Stretch
SRS: Septal Rebound Stretch
SSI: Systolic Stretch Index

## Availability of data and materials

The datasets generated and/or analyzed during the current study, as well as code needed to reproduce all aspects of the current study, are available in the Figshare repository, doi:10.6084/m9.figshare.15155596. The most recent version of the analysis code is available in the Github repository, https://github.com/dloewenstein/dillacs-study.

## Ethics approval and consent to participate

The study was approved by the local human subject research ethics committee at each site, and all subjects either provided written informed consent or were included following a retrospective waiver of informed consent provided by the local ethics committee.

## Authors’ contributions

DEL: conception and design of the work, data acquisition and analysis, interpretation of data, drafting the work. BW: data acquisition and analysis, interpretation of data, substantively revised the work. EH: data analysis, interpretation of data, substantively revised the work. JA: data acquisition, substantively revised the work. IK: design of the work, data acquisition, interpretation of data, substantively revised the work. RN: data acquisition, interpretation of data, substantively revised the work. EBS: data acquisition, substantively revised the work. PS: data acquisition, substantively revised the work. AS: data acquisition, substantively revised the work. DGS: design of the work, substantively revised the work. RJK: data acquisition, substantively revised the work. BDA: conception and design of the work, interpretation of data, drafting the work. MU: conception and design of the work, interpretation of data, drafting the work. All authors read and approved the final manuscript.

